# Buprenorphine treatment of opioid dependence: analysis of individual patient data

**DOI:** 10.1101/2020.03.18.20038430

**Authors:** Andrew W Bergen, James W Baurley, Carolyn M Ervin, Christopher S McMahan, Joe Bible, Randall S Stafford, Seshadri C Mudumbai, Andrew J Saxon

## Abstract

**Background:** The efficacy and safety of buprenorphine alone and in combination with naloxone for treatment of opioid dependence were evaluated in Federally-sponsored randomized clinical trials. Meta-analysis of pooled individual participant data provides an opportunity to identify multiple predictors of buprenorphine treatment outcome.

**Methods:** We selected six buprenorphine efficacy and safety trials from NIDA’s Data Share database for analysis. Treatment, sociodemographic, and drug use history variable domains were systematically harmonized and included in analysis. After exclusions, 3,022 participants randomized or enrolled in buprenorphine treatment for opioid dependence (mean (SD) age 36.1 (9.8) years, 33% female, 66% White, 16% Hispanic, 14% Black), were analyzed using a generalized linear mixed model with time-weighted treatment variables and participant covariates. We defined positive urinalysis or self-reported lapse as the primary outcome.

**Results:** Four treatment variables were significantly associated (*p* < 0.001) with lapse. Time-weighted dose and time-weighted adaptive dose had greater estimated effects than time-in-trial and time-weighted clinic visit. All treatment variables were novel predictors of lapse.

**Conclusions:** In a large cohort of trial participants treated with buprenorphine and behavioral counseling for opioid dependence, we identified and ranked four novel treatment factors reflecting components of buprenorphine dose, clinical provider engagement and patient engagement. Additional research to explore the effects of pharmacologic and non-pharmacologic treatment factors, and to explore relations with provider and patient factors will help our understanding of buprenorphine treatment outcomes. Continued analyses of publicly available data will extend discovery and support development of personalized opioid use disorder treatments.

**Highlights (3 to 5 bullet point max 85 characters each including spaces):**

- Treatment and participant variables were harmonized in six buprenorphine trials
- Time-weighted treatment variables were used in a random effects mixed model of lapse
- Buprenorphine dose and three clinical interactions were protective against lapse
- Support of protective treatment factors may improve buprenorphine treatment success

## 1. Introduction

In 2018, over two million adolescents and adults in the United States were estimated to have a opioid use disorder (OUD), and over 10 million were estimated to have misused prescription pain relievers and/or used heroin (Center for Behavioral Health Statistics and Quality 2019). Medication Assisted Treatment for OUD (MOUD) in the United States includes three FDA-approved medications combined with psychosocial and supportive therapies (Volkow et al. 2014). The medications bind to the mu opioid receptor (MOR), and include the full MOR agonist methadone (approved in 1972), the full MOR antagonist naltrexone (1984), and the partial MOR agonist buprenorphine, alone or in combination with the pure MOR antagonist, naloxone (both 2002). The approval of buprenorphine and buprenorphine-naloxone for MOUD in the United States represented decades of effort involving clinicians, scientists, and policy-makers in public agencies, medical institutes, pharmaceutical companies, and treatment clinics (Campbell and Lovell 2012).

Clinical researchers in the Addiction Research Center of NIDA and the Behavioral Pharmacology Research Unit of Johns Hopkins University performed early clinical studies and trials of buprenorphine (Bickel et al. 1988; Jasinski et al. 1978; Johnson et al. 1992, 1995). NIDA-supported research addressed a wide range of efficacy, safety and implementation issues in buprenorphine and buprenorphine-naloxone treatment trials in community-based clinics, coordinated by the VA’s Cooperative Studies Program and NIDA’s Clinical Trials Network. These trials were critical to translate and clinically validate buprenorphine MOUD as envisioned by the OUD treatment community (Ling et al. 2010). While significant restrictions remain, legislative, policy and regulatory changes were necessary to enable physicians to treat patients with buprenorphine in the office-based setting (McCance-Katz 2004).

Clinical practice guidelines on MOUD provide guidance on a wide range of patient, clinical assessment, MOUD treatment, concomittant treatment and co-morbid factors (Consensus and Expert Panelists 2004; Kampman and Jarvis 2015; Expert Panelists, Scientific Reviewers and Field Reviewers 2018). Buprenorphine MOUD guidelines emphasize the importance of clinical judgement with respect to buprenorphine dosing, and of concomitant psychosocial treatment (Fiellin et al. 2004; McCance-Katz 2004). The dosing, monitoring and duration of buprenorphine treatment remains a clinically complex area due to individual patient clinical and social factors (Farmer et al. 2015; Greenwald et al. 2014; Schuckit 2016). We analyzed publicly available individual participant data from six efficacy and safety trials of buprenorphine maintenance treatment with detailed logs of buprenorphine dosings and opioid use lapses. We harmonized variables and developed a generalized linear mixed model (GLMM) approach with novel conceptualizations of treatment variables to model the longitudinal nature of treatment effects. We identify and rank multiple treatment effects which emphasize the importance of buprenorphine dosing and clinical engagement, and provide a novel perspective on the complexity of factors influencing buprenorphine treatment

## 2. Material and Methods

### 2.1. Ethical Approval

Informed consent was obtained from all individual participants by the study coordinators and clinical investigators of the treatment trials. Trial data were systematically deidentified; detailed deidentification notes are available for each trial from datashare.nida.nih.gov.

### 2.2. Buprenorphine Treatment Trials

Clinical trial data are available from NIDA’s Division of Therapeutics and Medication Consequences (DTMC) and the Clinical Trials Network (CTN) at NIDA’s Data Share database (datashare.nida.nih.gov). In 2017, we used the keyword “opiate” to search the Data Share site, and identified 10 efficacy and safety trials involving detoxification or maintenance treatment of DSM-IV opioid dependence (**Table S1**). After review of trial treatment goals and outcomes, and data dictionary and trial data concordance, we selected six efficacy and safety trials focused on buprenorphine maintenance treatment for analysis (**Table 1** and **Table S2**). Two trials came to our attention after these analyses began (CTN-0051 and CTN-0010) and were not included.

**Table 1:**
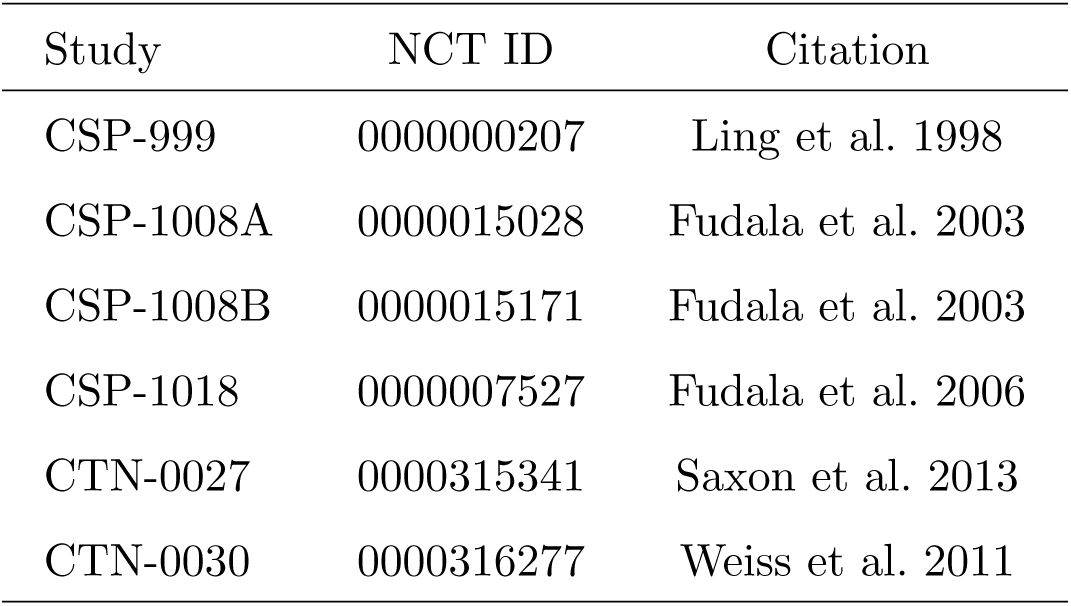
Treatment Trials Selected for Analysis

| Study | NCT ID | Citation |
| --- | --- | --- |
| CSP-999 | 0000000207 | Ling et al. 1998 |
| CSP-1008A | 0000015028 | Fudala et al. 2003 |
| CSP-1008B | 0000015171 | Fudala et al. 2003 |
| CSP-1018 | 0000007527 | Fudala et al. 2006 |
| CTN-0027 | 0000315341 | Saxon et al. 2013 |
| CTN-0030 | 0000316277 | Weiss et al. 2011 |

### 2.3. Literature Searches

We searched for and reviewed primary and secondary literature on the six trials to assess the type and design of published data analyses, using datashare.nida.nih.gov, clinicaltrials.gov, ctndisseminationlibrary.org, and ncbi.nlm.nih.gov/pubmed.

### 2.4. Eligibility Criteria and Trial Design

The eligibility criteria and design details of the six trials are provided in **Tables S3-S6**.

#### 2.4.1. Inclusion and Exclusion Criteria

Inclusion criteria for all trials (**Table S3**) include current opioid dependence by DSM-IV criteria (one trial required current dependence on prescription opioids), both males and females (excluding pregnant or breastfeeding females), with a common (four trials) minimum age of 18 years of age. One trial evaluated a small number of 15-18 year old individuals with opioid dependence for detoxification with continued maintenance treatment determined by clinical judgement, while the earliest trial admitted participants aged 21 to 50 years only. Participants were required to be in good physical health or in the care of a physician, to be agreeable with study conditions and able to give informed consent.

Exclusion criteria (**Tables S4 and S5**) were more numerous, but consistently excluded individuals with any acute medical condition that would make participation medically hazardous in the clinical judgement of the study investigator/physician, any current substance use dependence diagnosis excluding opiate, caffeine and nicotine dependence, expected inability to complete the study and refusal to use birth control if of childbearing potential. In addition, five of six trials had exclusions based on elevated liver enzymes, or recent participation in a methadone maintenance treatment program. Three trials had exclusions for: current severe psychiatric conditions; current alcohol, sedative or stimulant dependence requiring immediate medical attention; recent investigational drug study participation; and, recent or discontinued treatment with opioid agonists or naltrexone for opioid dependence.

#### 2.4.2. Trial Treatment Design

Treatment designs of the nine phases in six trials are annotated in **Table S6**. Four trials were randomized parallel designs. Protocols identify buprenorphine treatment safety, efficacy, both, or counseling efficacy as design objectives. Maximum treatment length was 12 months (two trials had extensions), buprenorphine/naloxone combination therapy was the most common pharmacotherapy, and flexible buprenorphine dosing based on clinical judgement was the most common treatment protocol (four, seven and seven trial phases, respectively). Double-blind randomized dose of active or inactive treatment occurred in two phases. Two trials provided one hour per week of psychosocial counseling, two provided 0.5 hours, one compared protocols of 0.35 *vs* 1.25 hours, and one relied upon usual clinic practice. Urinalysis or self-reported lapse (one trial) was a primary outcome in four trials, and a secondary outcome in two trials.

### 2.5. Trial Variable Harmonization

We performed variable harmonization using publicly available trial data dictionaries, protocols, data collection forms, and patient-level data, in a systematic step-wise fashion. We performed a review of the protocols and summarized the design, treatments, and outcomes. We grouped variables by domain and cataloged the information available from each trial. Domains included sociodemographics, methadone treatment history, drug use history, route of administration, substance use disorder and psychiatric diagnoses, buprenorphine dosage, clinical visits, and opiate usage (self-reported and urinalysis). We matched the variables across the trials and for select variables, inspected the coding, labels, and distributions in each trial. We created new harmonized variables across trials. Consultation with clinical experts occurred twice during harmonization.

### 2.6. Modeling Strategy and Outcome

Our analysis strategy focused on the evaluation of buprenorphine MOUD factors on use lapses. From the 4,853 individuals screened in the six trials (**Table S7**), we selected individuals randomized or enrolled into buprenorphine treatment including those randomized to placebo, and excluded participants randomized to methadone treatment or not randomized or enrolled into treatment. Urinalysis or self-reported lapse was selected as the outcome variable. We were interested in the recent, time-weighted effects of treatment variables on urinalysis or self-reported lapse (see **Section 2.8**).

### 2.7. Selection of Baseline Patient Variables as Covariates

We selected established sociodemographic and drug use history variables as covariates for this analysis. We report distributions but not model effect sizes of sociodemographic and drug use history covariates.

### 2.8. Development of Treatment Variables

We harmonized treatment variables: dose (mg buprenorphine/day), adaptive dose (whether a dose was the result of a clinical provider’s judgement *vs* a fixed protocol dose), clinic visit (whether the dose was self-administered in the clinic under observation *vs* taken home and self-administered), and time-in-trial (days in treatment) across trials. We conceptualized a model where treatment effects on lapse depend on treatment prior to the day of the urinalysis screen or self-report. We were guided by the extended half-life of sublingually- administered buprenorphine (Kuhlman et al. 1996), the hypothesis that clinical provider and patient engagement has positive effects on treatment outcomes, and the hypothesis that “clinically supervised administration may promote therapeutic engagement” (Saulle et al. 2017). We implemented this using a basic time-weighted model using three derived variables; Time Weighted Dose (TWD), Time Weighted Adaptive Dose (TWAD), and Time Weighted Clinic Visit (TWCV). At the *j*th visit occurring at time *t*_*j*_, the time-weighted variables are constructed using all values and weights of (1*/*2)^*t*^, for *t* = 0, …, *t*_*j*_ − 2, with the more recent values of the variables being given the higher weights. That is, the time-weighted variables take into account prior treatment effects, discounted by 50% for each dose period. For example, let *D*_*it*_ denote the dose level taken by the *i*th subject at time *t*, the time-weighted dose at *tj* would be 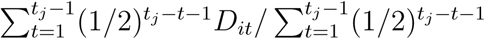. Note also that average time-weighting limits the maximal value of both quantitative and categorical variables. Time-in-trial was used as the fourth treatment variable; this variable is conceptualized to represent the effects of patient persistence in treatment and therapy effects received by the patient over time.

### 2.9. Selection of Candidate Risk Factors

From 103 harmonized variables across six trials, we selected 79 candidate factors to be examined. After filtering out candidate predictors with a missingness greater than 25%, we were left with 22 candidate risk factors (4 treatment, 9 sociodemographic and 9 drug use history factors with a total of 58 variable effects to be estimated) and 3,022 participants (**Tables 2, S8 and S9**). We imputed missing entries using the regularized iterative factorial analysis for mixed data (qualitative and quantitative variables) algorithm (Audigier et al. 2016), implemented by the imputeFAMD function in the missMDA R package. Finally, for computational stability, all quantitative variables were standardized to have a mean of zero and a standard deviation of one.

**Table 2:** Treatment and Outcome Characteristics

| Variable | Total N | Mean | Median | Range |
| --- | --- | --- | --- | --- |
| TWD<sup>α</sup> | 55,739 | 11.94 | 12 | 0-32 |
| TWAD<sup>β</sup> | 55,739 | 0.76 | 1.00 | 0-1 |
| TWCV<sup>γ</sup> | 55,739 | 0.34 | 0.03 | 0-1 |
| TinT (days)<sup>δ</sup> | 55,739 | 112.69 | 87 | 1-527 |
| Urinalysis (Yes=1)<sup>ε</sup> | 55,739 | 0.41 | 0 | 0,1 |

<sup>α</sup>TWD=Time-Weighted Dose. <sup>β</sup>TWAD=Time-Weighted Adaptive Dose. <sup>γ</sup>TWCV=Time-Weighted Clinic Visit. <sup>δ</sup>TinT=Time-in-Trial. <sup>ε</sup>Urinalysis screen or self-report.

### 2.10. Generalized Linear Mixed Model

Define *Y*_*ij*_, for *j* = 1, …, *n*_*i*_ and *i* = 1, …, *m*, to be a binary indicator, such that *Y*_*ij*_ = 1 denotes the event that the *i*th individual tests positively for opioids during the *j*th visit and *Y*_*ij*_ = 0 otherwise. To relate the covariates to the binary response, we posit the following generalized linear mixed model

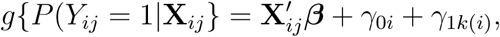

where *g*(*·*) is the logistic link, **X**_*ij*_ is a vector of covariate information (e.g., see **Tables S7 and S8**), *γ*_0*i*_ is a subject specific random effect specified to account for the longitudinal nature of the data, *γ*_1*k*(*i*)_ = *γ*_1*k*_ if the the *i*th subject is a part of the *k*th trial, for *k* = 1, …, *K*, and *γ*_1*k*_ is a random effect specified to account for the heterogeneity across trials. Herein, the random effects distributions were taken to be

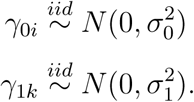

Note, here the random effects are specified to be independent given their nesting. Model fitting was completed via PROC MIXED in SAS version 9.4 (Cary, NC).

### 22.11. Assessment of Results

A p-value < 0.001 was considered statistically significant based on the following logic: 58 variable effect estimates were estimated in 22 treatment, demographic and drug use domains; a Bonferroni correction (0.05*/*58 = 0.00086) is conservative, thus, we chose *α* < 0.001, slightly less conservative. Nominal significance was declared for effect size estimates with a p-value < 0.01. Searches of treatment variable-related terms were conducted to assess novelty as described in **Table S12**. The literature was reviewed to provide support for external validity.

## 3. Results

**Tables S8, S9 and 2** provide sociodemographic, drug use history, and treatment and outcome variable characteristics. **Figures 1 - 3** and accompanying text describe treatment and outcome variables in the analysis dataset, while **Table 3** provides effect estimates of treatment and selected anthropometric variables from our longitudinal analysis.

**Table 3:**
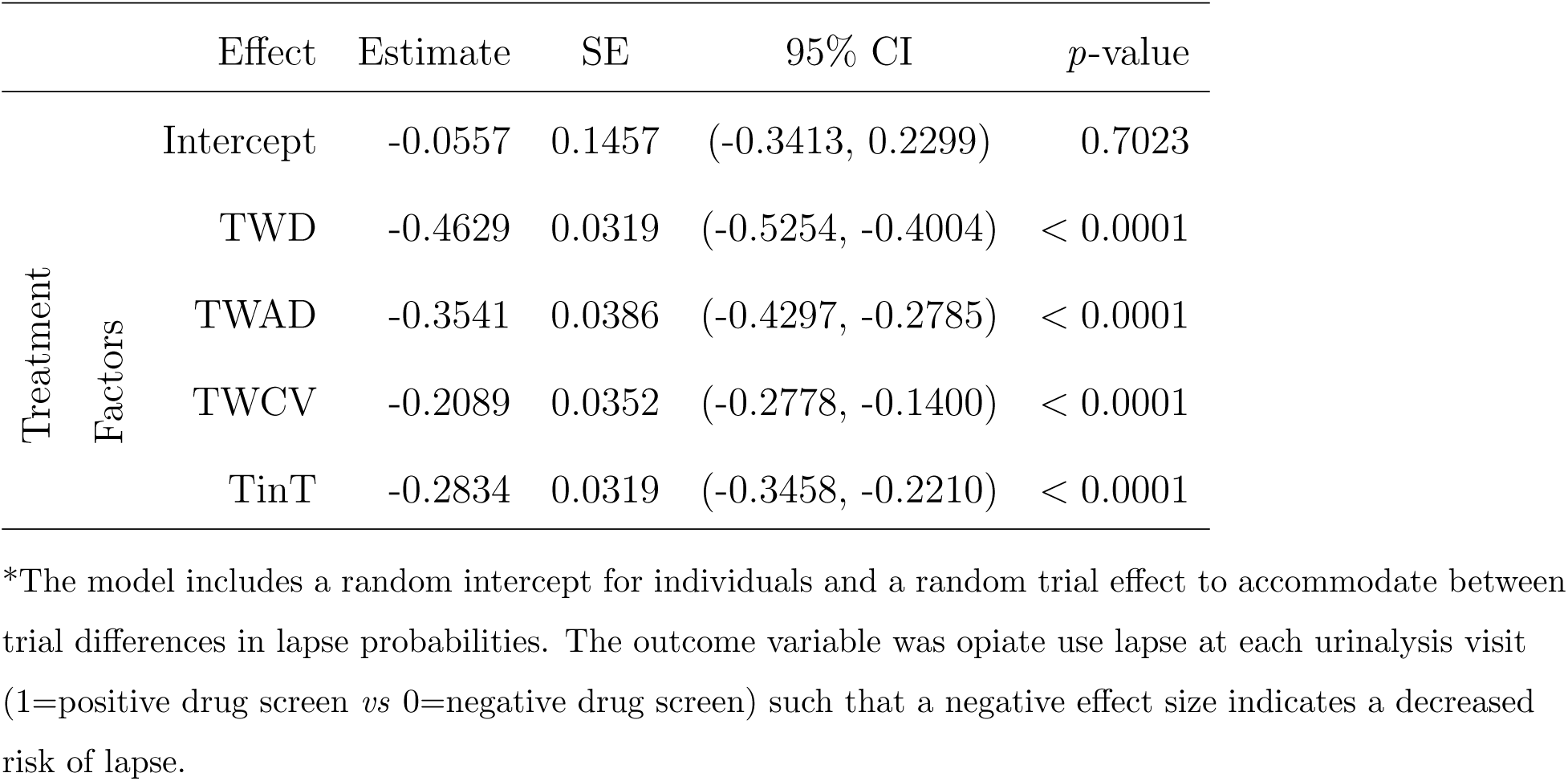
Fixed effect parameter estimates for the GLLM of lapse*

**Figure 1:**
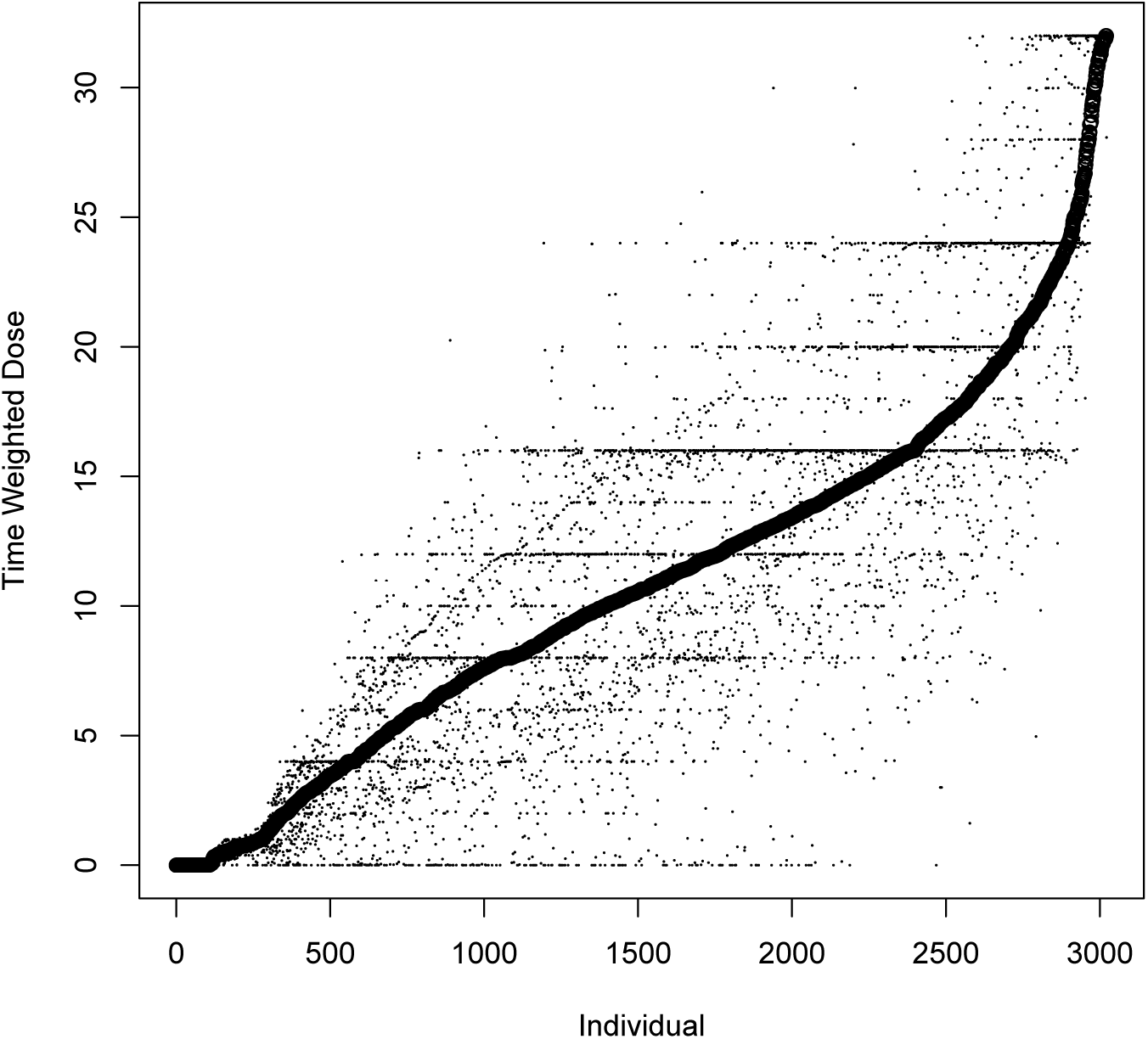
Mean, as well as the 25% and 75% quantiles, of the time-weighted dose of the 3,021 individuals, i.e., effectively subject specific box-plots of time-weighted dose. Individuals (on the x-axis) are orderd from smallest to largest based on time-weighted mean dose.

### 3.1. Participant Characteristics

#### 3.1.1. Sociodemographics

Of 3,022 individuals from six trials included in the mixed model analysis (**Table S8**), the mean (standard deviation) age were 36.1 (9.8) years, 33% of individuals were female, while 66% were White, 16% Hispanic, and 14% were Black. Nearly half of the sample had a high school education. Two-thirds of the sample had employment histories of skilled manual work, never gainfully employed, or semi-skilled manual work. Most individuals were in full time work. Two-thirds of the sample were married or had never married, while nearly all lived with a partner, family or friends.

#### 3.1.2. Drug Use History

All trial participants fulfilled opiate or prescription opioid dependence criteria. Nearly 80% of the sample reported a heroin use history (**Table S9**). The mean and standard deviation of years of opiate/opioid use were just over 8 years. The most common modes of opiate or opioid use were intravenous (56%) and nasal (snorting) (36%). Non-opiate/opioid drug use history, excluding tobacco use history, is available in all trials and was common. Nearly two thirds of the sample admitted to a marijuana, alcohol or cocaine use history, one third to a tranquilizer use history, one quarter to a methamphetamine use history, and one-sixth to a phencyclidine use history.

#### 3.1.3. Treatment and Outcome Characteristics

Descriptive statistics and explanatory text for the four unstandardized treatment variables and the outcome variable are in **Table 2**. A total of 479,151 dose records were used to construct the three time-weighted continuous derived variables at the 55,739 urinalysis or self-report records in the 3,022 participants included in the analysis. For clarity, a summary of the distribution of unstandardized mean time-weighted dose is plotted in **Figure 1**; i.e., each point on the x-axis represents a participant and the y-axis presents the 25%, the mean and the 75% of the time-weighted dose values of each participant. **Figure 1** demonstrates the variability of time-weighted dose across and within participants. Note that in **Figure 1** we excluded one individual with a dosage record consistent with induction with buprenorphine, then transition to methadone [8 12 16 24 0 20 30 40 50 60 70 80 90]. An analysis excluding this individual was conducted and resulted in no appreciable differences in effect estimates and no changes in significant variables (data not shown). Time-in-trial is a continuous variable recording the number of days in treatment for each of the 55,739 urinalysis results (Urinalysis) included in the analysis.

The descriptive statistics of the treatment variables and the outcome provides immediate insight into the characteristics of treatment and lapse over the entire analytic sample. **Figure 1** and **Table 2** provide two perspectives on the distribution of time-weighted dose. The mean time-weighted adaptive dose reflects seven trial phases with flexible dosing and two trial phases with fixed doses (see **Table S5**). The mean and median values of time-weighted clinic visit suggest that most doses were self-administered at home, with a minority of patients self-administering more doses in the clinic. The mean Urinalysis value is similar to the inverse of a Total Effectiveness Score expressed as a percentile (Ling et al. 1997), here calculated over all participants. The mean of Urinalysis suggests that about two-fifths of all records on lapse report illicit opioid use. Comparing the mean and median of the Time-in-trial variable gives a sense of its distribution, i.e., the mean was a week less than four months, while the median was about three months.

We plot the time-weighted doses of a single randomly selected participant in **Figure 2** to illustrate the pattern of variation. The numerical daily dose, urinalysis days and time-weighted dose data from this individual are found in **Table S10**. E.g., this individual missed doses on days 40 and 41, days 67 and 72, and days 86 and 93. The missed doses have the effect of reducing the time-weighted dose at days 41, 72, and 93. The minimum time-weighted dose (16.00 at day 41) is the result of the missed doses on days 40 and 41, while the next minimum time-weighted dose (23.63 at day 171) is due to the tapering doses at the end of the trial. The maximum time-weighted dose is 32.00 at day 35, i.e., it is reached early in the trial before missing doses are recorded, however there are many time-weighted doses later in the trial that approach 32 (see **Table S10**).

**Figure 2:**
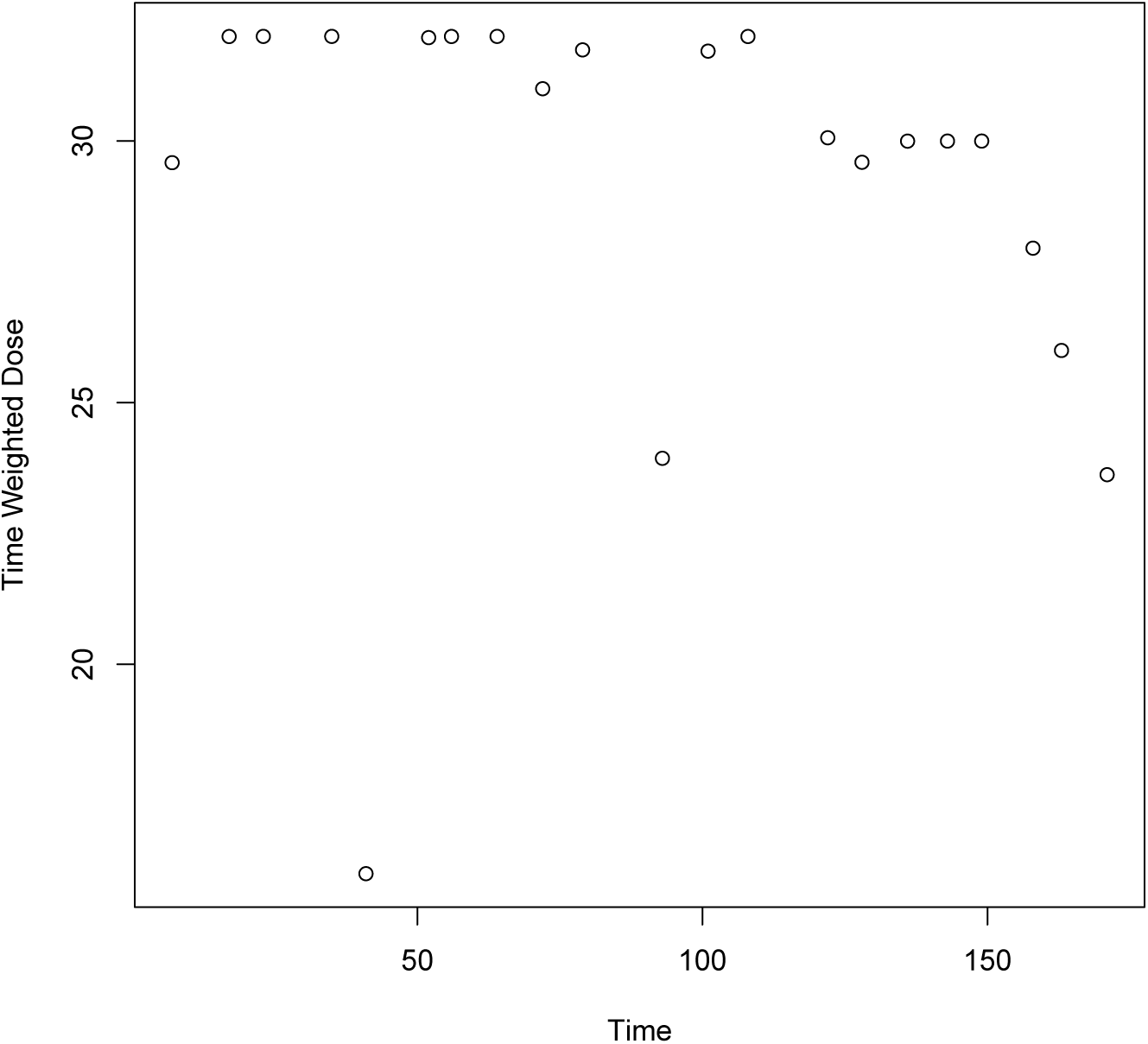
Time weighted dose of one individual trial participant (see **Table S9**).

Urinalysis tests per individual by mean time-weighted dose are plotted in **Figure 3**. It is apparent that those individuals prescribed lower doses are more likely to have fewer urinalysis tests than individuals prescribed with higher doses (see the lines of points on the lower left *vs* the middle right of the plot). However, there are some individuals who are prescribed low doses of buprenorphine and remain in treatment for a full year of urinalysis tests (see the line at the top left of the figure at 52 urinalysis tests).

**Figure 3:**
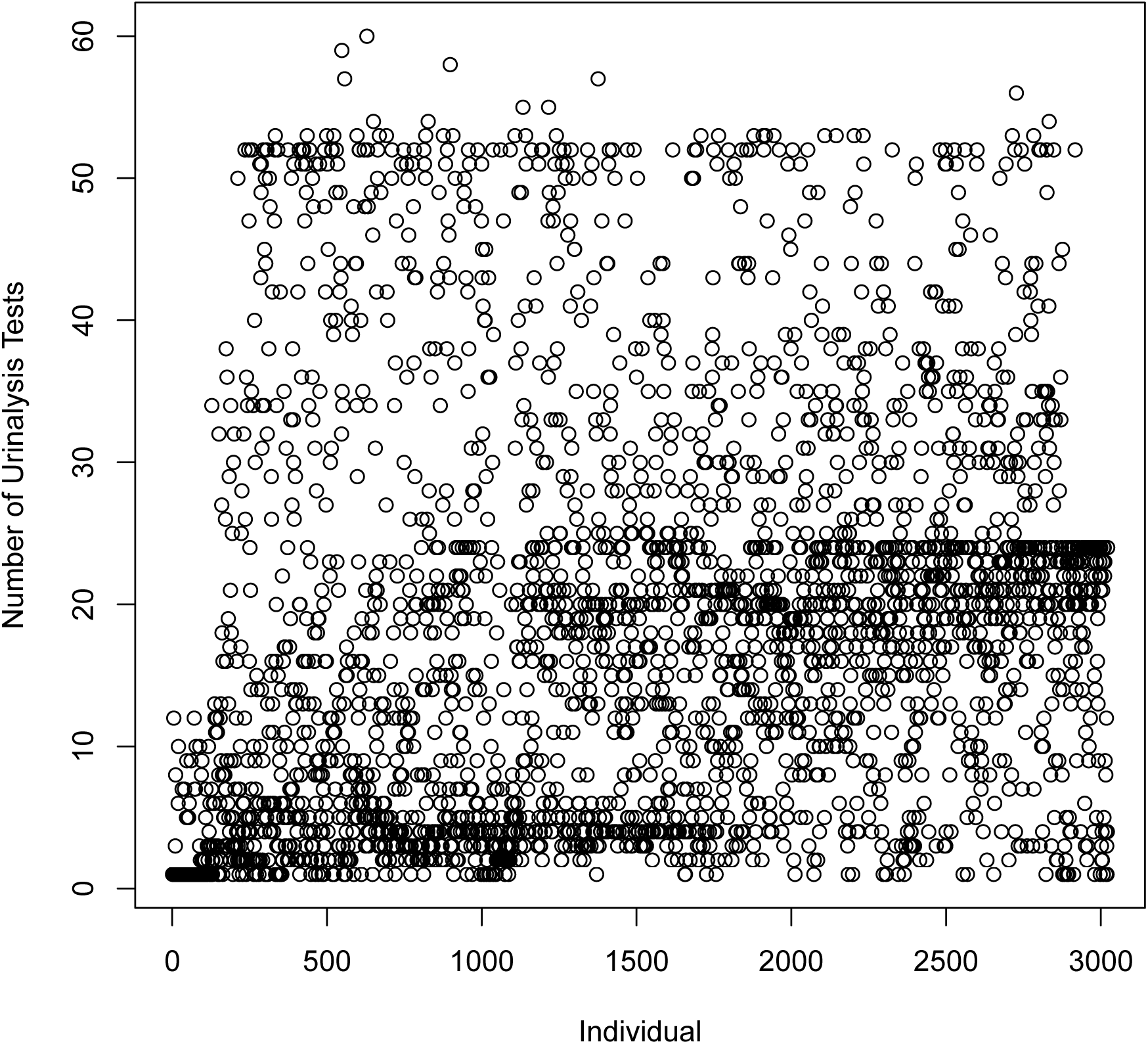
The number of urinalysis records by individual ranked by mean TWD.

### 3.2. Results from GLMM Analysis

We identified statistically significant (*p* < .001) protective effects against lapse associated with each of the four novel treatment variables (**Table 3**).

### 3.3. Prior Published Analyses of the Six Trials

All six trials are represented by at least one individual participant data analysis, five are represented by summary data analyses in the meta-analytic or systematic review literature, and two are represented by multiple secondary data analyses of individual participant data (**Table S11**).

## 4. Discussion

We investigated the influence of multiple treatment variables on lapse, using publicly available individual participant data from NIDA-sponsored buprenorphine efficacy and safety trials. With respect to buprenorphine MOUD treatment effects comparable to the questions we asked in our analysis, meta-analyses have identified buprenorphine to be superior to placebo with respect to illicit opioid use (positive urinalysis) at doses ≥ 16 milligrams (Mattick et al. 2014), but failed to judge whether supervised dosing *vs* dispensing of buprenorphine to be consumed away from the dispensing point is more effective with respect to abstinence or retention (Saulle et al. 2017). In our analyses, we address effects of buprenorphine dose (Mattick et al. 2014) and of supervised dosing (Saulle et al. 2017), in addition to adaptive dosing and time-in-treatment, in relation to lapse. We discuss validity, strengths and limitations, and clinical and research implications of our findings below.

### 4.1. Treatment Factor Findings

Our novel conceptualizations of time-weighted buprenorphine dose, adaptive dose, and clinic dose, and our novel use of time-in-trial, were found to be statistically significant protective treatment factors.

Time-weighted dose of buprenorphine was the most protective treatment factor, reflecting the efficacy of buprenorphine (Ling et al. 2010). The formal conceptualization of time-weighted dose is a initial model for the recent effects of buprenorphine dose, including recent increases or decreases in daily dose, and of missed doses.

The effects of time-weighted adaptive dose on lapse, the second ranked treatment factor, reflect the protective association of flexible dosing (physician interaction with the patient and physician clinical judgement), while dose effects are accounted for by time-weighted dose.

The third ranked treatment variable, time-in-treatment, is usually analyzed as an outcome variable, i.e., time-in-treatment is usually conceptualized as an effect of treatment, which also predicts better long-term outcomes (Timko et al. 2016). In our analysis, we used time-in-treatment as a predictor of lapse, a novel application. Analyses of Massachusetts Medicaid administrative data identified opioid agonist treatment length as significantly protective against relapse (Clark et al. 2015), suggesting our finding is a related example.

Effects of the fourth ranked time-weighted clinical dose variable reflect patient interaction with the treatment clinic environment and clinical staff while self-administering a dose, i.e., the beneficial effect of the clinical environment, and of clinical staff-patient engagement.

### 4.2. Limitations

We focused harmonization efforts on variables with data available across most trials. Harmonization was limited by different eligibility criteria, some missing data dictionaries and discordant or incomplete coding of some variables. Therefore, we did not perform subset analyses to include important covariates such as psychiatric comorbidity or chronic pain, common among patients with OUDs (Katz et al. 2013; Lusted et al. 2013). Such analyses would address these participant factors in buprenorphine MOUD that complicate functional restoration of the patient.

### 4.3. Treatment Implications

The four novel treatment factors that arise from pharmacological sources and non-pharmacologic sources illustrate the complexity of factors that lead to reduction of illicit opioid use. Clinical implications of the novel treatment variable effect estimates are both quantitative and qualitative. Ranked by estimated effect size:

- Buprenorphine dose is the most important treatment factor in buprenorphine MOUD. The mean (standard deviation, SD) buprenorphine dose and mean maximum dose (SD) of reported in 2015-2018 by N=1,105 physicians treating OUD patients with buprenorphine-naloxone for stable patients were 13.4 (4.2) and 21.5 (6.2) milligrams (Knudsen et al. 2019). Thus recent practice covers the entire range of recommended and appropriate buprenorphine daily doses (Expert Panelists, Scientific Reviewers and Field Reviewers 2018). In the clinical trial sample we analyzed, time-weighted dose covers the entire range of recommended and appropriate buprenorphine daily doses with mean, median and interquartile range of mean time-weighted dose of 11.94, 12, and 8 to 14 milligrams (IQR is Bergen’s estimate from Figure 1), respectively. We acknowledge differences in patient characteristics and dosing protocols between the clinical trial sample and current office-based practice.
- The protective effect of adaptive dosing and the dose distribution (**Figure 1**) suggests that buprenorphine MOUD is highly personalized therapy (Greenwald et al. 2014). In a national analysis of responses to 16 practice vignettes by N=1,105 physicians prescribing buprenorphine-naloxone, an upward adjustment of daily dose was the suggestive response to one patient vignette (“8 mg daily dose, usually positive urinalysis with euphoria from opioid use, a history of intravenous use and fresh tracks, rescheduled appointments at last minute, adherent to treatment, private insurance and bi-weekly office visits”), however, most (12/16) patient vignettes elicited scores suggesting no change of dose (Knudsen et al. 2019). This possible test of external validity suggests that adaptive dosing may be utilized less frequently in current office-based practice compared to the clinical trial sample. We acknowledge there are differences in patient characteristics, dosing protocols and the measures of adaptive dosing between the clinical trial sample and the office-based practice survey.
- Time-in-treatment may represent *both* patient motivation, skills and persistence in treatment (Strong et al. 2012), *and* treatment therapy effects delivered over time, in the apparently simple construct of days in treatment. The effect size of time-in-treatment on lapse is less than adaptive dosing and greater than clinic visits (**Table 3**). This suggests that beneficial buprenorphine treatment effects of successful efforts to retain patients in treatment may lie between the beneficial effects of successfully implemented adaptive dosing and clinical observation of self-administered doses.
- The clinic visit factor has the lowest effect size of the four treatment factors analyzed. However, in Knudsen *et al*, physicians reported that they were likely to increase the frequency of office visits in response to all 16 patient vignettes, while the individual criterion with largest relative importance score associated with increasing office visits (after office visits itself) was “positive urinalysis with euphoria from opioid use”. In the same study, about three-quarters of physicians reported monthly office visits for stable patients with no ongoing substance use, and weekly or biweekly office visits for patients showing signs of clinical instability. We acknowledge that there are differences in self-administration in the clinical setting (and other clinical setting practices) between the clinical trial sample and the office-based practice survey.

#### 4.3.1. Summary of Treatment Implications

Increasing physician-patient and practice-patient engagement resulting in more opportunities to evaluate patients, adapt buprenorphine dose, retain patients, and increase the frequency of office visits, should be possible, given that most waivered physicians are seeing fewer patients than permitted, and often have months with no buprenorphine treatment episodes (Thomas et al. 2017).

### 4.4. Research Implications

Each of the treatment factor findings suggests future research hypotheses that might be explored in this or similar datasets of individual patient data from buprenorphine MOUD. E.g., the time-weighted dose treatment finding may support use of the maximum induction dose recommended by clinical guidelines (Expert Panelists, Scientific Reviewers and Field Reviewers 2018), but more specific analyses of dose and lapse during induction are needed to support this specific hypothesis. Identification of more subtle relations between dose and lapse, e.g., by addiction severity (McLellan et al. 1980), specific comorbidities (Katz et al. 2013; Lusted et al. 2013), or demographic and clinical characteristics (Lapham et al. 2019) would help address dose and participant related complexities of buprenorphine MOUD.

Identification of engagement and patient factors that influence the factors associated with non-pharmacologic treatment metrics is another research priority. I.e., the three treatment findings after buprenorphine dose level include several clinical interactions. Each interaction could be further examined using physician or clinical staff characteristics, engagement metrics, and patient characteristics, to identify relations that might lead to improved understanding of treatment outcomes. An analysis challenge will be to locate datasets of buprenorphine MOUD with such detail.

Beyond these additional analyses of individual protective treatment factors lies the more complex effort of understanding which combination of protective treatment factors might result in more effective, long-term functional restoration. E.g., whether increasing the frequency of office visits or increasing dose (Knudsen et al. 2019), or whether combinations of two or more protective treatment factors will most effectively reduce positive urinalysis over the long term is a question for future factorial clinical research (Baker et al. 2017).

#### 4.4.1. Summary of Research Implications

Additional public data sharing and analysis of buprenorphine treatment trials and administrative cohorts will contribute to a greater understanding of factors that might be utilized in translational buprenorphine MOUD research. Future translational research would support efforts to treat the majority of individuals with OUD who are not in MOUD treatment.

## 5. Conclusion

We present evidence from analysis of publicly available data from 3,022 individuals randomized or enrolled into buprenorphine maintenance treatment for DSM-IV opioid dependence. We identify, rank and interpret significant effects of four novel treatment variables. Treatment variable findings support four clinical implications (two of which are consistent with existing guidance) that may reduce illicit opioid use in buprenorphine treatment: a) adequate dosing, b) physician tailoring of buprenorphine dose, c) retaining patients in treatment, and d) patient self-administration of buprenorphine in the clinical setting.

## Data Availability

Clinical trial data are available from NIDA's Division of Therapeutics and Medication Consequences (DTMC) and the Clinical Trials Network (CTN) at NIDA's Data Share database (datashare.nida.nih.gov).

https://datashare.nida.nih.gov/

## Author Disclosures

### Contributors

Drs. Bergen and Baurley were responsible for obtaining funds to support the study. Drs. Bergen, Baurley, Ervin, and McMahan designed the study. Dr. Baurley developed the trial database and coded the treatment variables. Drs. Bergen and Ervin harmonized the variables. Drs. Saxon and Mudumbai provided clinical consultation on variable harmonization. Drs. Baurley and McMahan designed the analysis pipeline. Drs. McMahan and Bible performed variable imputation and modeling of variable effects on lapse. Drs. Bergen, Baurley, McMahan, Mudumbai, Stafford and Saxon wrote the ms. All authors reviewed the ms, agreed with its contents and agreed to its submission.

## Acknowledgements

The information reported here results from secondary analyses of data from clinical trials conducted by the National Institute on Drug Abuse (NIDA). Specifically, data from the following trials were included: CSP-999, “A Multicenter Clinical Trial of Buprenorphine in Treatment of Opiate Dependence”; CSP-1008A, “A Multicenter Efficacy/Safety Trial of Buprenorphine/Naloxone for the Treatment of Opiate Dependence”; CSP-1008B, “A Multicenter Safety Trial of Buprenorphine/Naloxone for the Treatment of Opiate Dependence”; CSP-1018, “A Multicenter Safety Trial of Buprenorphine/Naloxone for the Treatment of Opiate Dependence”; CTN-0027, “Starting Treatment with Agonist Replacement Therapies (START)”; and CTN-0030, “A Two-Phase Randomized Controlled Clinical Trial of Buprenorphine/Naloxone Treatment Plus Individual Drug Counseling for Opioid Analgesic Dependence”. NIDA databases and information are available at http://datashare.nida.nih.gov.

